# Clinical Outcomes of Switching vs. Continuing Direct Oral Anticoagulants (DOACs) After Ischemic Stroke in Patients with Atrial Fibrillation in the US

**DOI:** 10.64898/2026.07.06.26357356

**Authors:** Chiang Jia-Herng, Alvaro Alonso

## Abstract

**Background:** Clinical outcomes of switching versus continuing direct oral anticoagulant (DOAC) among atrial fibrillation (AF) patients who experienced an ischemic stroke despite receiving DOAC therapy are uncertain.

**Methods:** We included patients with AF who were hospitalized for ischemic stroke (index stroke) between January 1, 2016, and June 30, 2022, while receiving DOAC therapy and who resumed DOAC within 90 days after discharge in the Merative MarketScan Commercial and Medicare databases. Patients were classified as DOAC-switched or DOAC-continued according to whether the DOAC agent changed or remained the same after the index stroke; secondary analyses considered individual DOACs. The primary outcome was recurrent ischemic stroke; secondary outcomes included major bleeding and a composite outcome (bleeding or ischemic stroke). Propensity score–based overlap weighting and weighted Cox models were used to estimate adjusted hazard ratios (aHRs).

**Results:** A total of 1175 patients were eligible for the study, of whom 970 (82.6%) continued and 205 (17.4%) switched DOAC therapy. Comparing DOAC-switched to DOAC-continued was not significantly associated with recurrent ischemic stroke (aHR, 1.20; 95% CI, 0.63-2.30), major bleeding (aHR, 0.60; 95% CI, 0.21-1.72), or the composite outcome (aHR, 0.98; 95% CI, 0.56-1.70). However, among patients who received apixaban before stroke, switching to rivaroxaban was associated with a higher risk of recurrent ischemic stroke (aHR, 2.70; 95% CI, 1.05–6.95).

**Conclusions:** Overall, switching DOAC therapy after ischemic stroke was not associated with improved clinical outcomes. Switching from apixaban to rivaroxaban, however, could increase risk of recurrent ischemic stroke.

**Clinical Perspective:** *What Is New?:*

- Among patients with atrial fibrillation who experienced ischemic stroke while receiving DOAC therapy, switching to another DOAC was not associated with lower risks of recurrent ischemic stroke or major bleeding compared with continuing the same DOAC.
- Switching to a DOAC with a different mechanism of action or adding antiplatelet therapy was not associated with improved outcomes, whereas switching from apixaban to rivaroxaban was associated with a higher risk of recurrent ischemic stroke.

*What Are the Clinical Implications?:*

- These findings support current guideline recommendations against anticoagulant switching or antiplatelet addition after ischemic stroke during DOAC therapy, although individual DOAC selection may still influence recurrent stroke risk.

## Background

The prevalence of atrial fibrillation (AF) is increasing in the United States, with an estimated prevalence of at least 4.48%, corresponding to approximately 10.55 million affected adults at the end of 2019.^1^ AF is associated with a 2.3-fold increased risk of ischemic stroke.^2^ A large meta-analysis that combined population- and hospital-based studies from 1993 to 2017 worldwide reported that 22% of ischemic stroke were attributed to cardioembolism.^3^ Therefore, patients who are diagnosed with AF should undergo risk assessment for thromboembolic events using a validated clinical risk score, such as CHA2DS2-VASc, and receive appropriate antithrombotic therapy.^4^

Oral anticoagulants can prevent ischemic stroke in patients with AF but meanwhile increase the risk of bleeding. A meta-analysis of RCTs that evaluated different antithrombotic therapies among non-valvular AF patients found that warfarin, compared to placebo, reduced the risk of ischemic stroke by 67%. ^5^ Warfarin was used as a first-line therapy until direct oral anticoagulants (DOACs) came into practice in the early 2010s. A meta-analysis of RCTs comparing individual DOACs (apixaban, dabigatran, edoxaban, and rivaroxaban) with warfarin showed that DOAC was noninferior to warfarin in ischemic stroke prevention and major bleeding events among patients with AF. ^6^ Given the disadvantages of frequent monitoring of the INR to maintain a therapeutic range, more frequent drug interactions, and dietary restrictions of warfarin, current practice guidelines recommend DOACs over warfarin for thromboembolism prevention in AF patients with CHA2DS2-VASc score of ≥2 in men and ≥3 in women, in the absence of mechanical heart valves or moderate-to-severe mitral stenosis. ^4,7^

However, despite receiving appropriate DOACs, patients enrolled in DOAC trials experienced 0.92% - 1.25% annual rate of ischemic stroke and 0.08% - 0.09% annual rate of systemic embolism^8–10^ Potential causes include competing stroke mechanisms, such as large artery disease and small vessel disease; cardioembolism with insufficient anticoagulation, including nonadherence, inappropriate dosing, or low anticoagulant activity despite sufficient dosing; and cardioembolism despite sufficient anticoagulation. Among these, cardioembolism despite sufficient anticoagulation is the most common cause, accounting for 44.1% of the events.^11^ The anticoagulant strategies after ischemic stroke in these patients differ across neurologists. Common practices include switching to another DOAC, switching to warfarin, or adding an antiplatelet drug. Current AF guidelines state that for patients who experience an ischemic stroke while receiving oral anticoagulant therapy, and after assessment of non-cardioembolic causes, dosage, and adherence, adding antiplatelet therapy or switching anticoagulants is not recommended.^7^ However, the supporting studies for these recommendations are limited and lack long-term evaluation,^11–13^ Moreover, none of the previous studies focused exclusively on American populations. ^11–16^ This study aimed to address limitation of the current evidence by comparing the clinical outcomes of continuing versus switching DOAC among AF patients who experienced an ischemic stroke despite receiving DOAC therapy in the United States.

## Methods

### Study Design and Data Source

This is a retrospective cohort study. We used data from Merative MarketScan Commercial and Medicare databases. The MarketScan databases contain individual-level, de-identified longitudinal information on inpatient and outpatient medical claims, including diagnosis, procedures, and healthcare services, as well as outpatient prescription drug claims and enrollment records. These data capture healthcare utilization among insured employees and their dependents for active employees, early retirees, Consolidated Omnibus Budget Reconciliation Act (COBRA) continuers, and Medicare-eligible retirees enrolled in employer-provided Medicare Supplemental or Medicare Advantage plans. This study was reviewed and approved by the Emory University Institutional Review Board and was granted a waiver of informed consent.

### Study Subjects

We identified patients with AF who were hospitalized for ischemic stroke between January 1, 2016, and June 30, 2022, while receiving DOAC therapy, including apixaban, dabigatran, edoxaban, or rivaroxaban, and who subsequently resumed DOAC treatment after the ischemic stroke. This ischemic stroke event is thereafter referred to as the index stroke. The diagnosis of AF required the presence of International Classification of Diseases, Tenth Revision, Clinical Modification (ICD-10-CM) code I48.xx in 1 inpatient claim or 2 outpatients claims that were 7 to 365 days apart, with the AF diagnosis required to precede the index ischemic stroke. Ischemic stroke was defined by ICD-10-CM code I63.xx listed as the primary diagnosis in an inpatient claim. Patients were required to have at least one DOAC prescription within 180 days before ischemic stroke admission. Patients were excluded if they had mitral stenosis, mechanical valvular replacement, percutaneous or surgical left atrial appendage occlusion, left ventricular thrombus, atrial septal defect, active malignancy, polycythemia rubra vera, essential thrombocytosis, as defined by ICD-10-CM codes listed in the Supplementary Materials (Table S1). Only patients who resumed DOAC within 90 days after discharge from the ischemic stroke hospitalization were included in the study.

### Exposure

We compared the type of DOAC therapy prescribed before the index stroke with that prescribed after the index stroke within each patient, irrespective of dosage. Patients who received same type of DOAC before and after the index stroke were classified as DOAC-continued group, whereas those whose DOAC were changed after the index stroke were classified as DOAC-switched group.

### Outcomes

The primary outcome was recurrent ischemic stroke occurring after the index stroke. The secondary outcome was major bleeding, defined as bleeding events that resulted in hospitalization, including intracranial hemorrhage, gastrointestinal bleeding, and other bleeding sources defined by ICD-10-CM codes listed in the Supplementary Materials (Table S1). The composite outcome was defined as a combination of recurrent ischemic stroke and major bleeding.

### Follow-up Period

The date of the first DOAC prescription after the index stroke was defined as the index date. Follow-up was defined as the period from the index date to the occurrence of a study outcome, disenrollment from the insurance program, or the end of the study, June 30, 2022, whichever occurred first. Patients who experienced recurrent ischemic stroke before initiation of DOAC therapy after the index stroke were excluded since their exposure group could not be defined.

### Baseline Covariates

Variables that may influence the choice of DOAC strategy, the risk of study outcomes, or both were collected. These variables included sex, age at the start of follow-up, history of alcohol use, preexisting medical comorbidities, use of antiplatelet agents (aspirin, clopidogrel, or ticagrelor) within 180 days before the index stroke, and concomitant medication use after resumption of DOAC therapy.

Preexisting medical comorbidities, diagnosed before the start of follow-up, included heart failure, hypertension, diabetes mellitus, hyperlipidemia, coronary artery disease, peripheral artery disease, venous thromboembolism, chronic kidney disease, chronic liver disease, and anemia, defined by ICD-10-CM codes listed in the Supplementary Materials (Table S1).

Concomitant medication use was defined as prescriptions within 90 days after discharge from the index stroke hospitalization and included antiplatelet agents (aspirin, clopidogrel, or ticagrelor), CYP or P-gp modulators (rifampin, carbamazepine, phenytoin, phenobarbital, dronedarone, amiodarone, cyclosporine, and verapamil), NSAIDs, proton pump inhibitors, and statins.

### Statistical Analyses

Baseline characteristics were presented as means with standard deviations (SDs) and counts with percentages for categorical variables. To balance the baseline covariates between the DOAC-same and DOAC-switch groups, we applied propensity score-based overlap weighting.^17^ Propensity scores were estimated using a multivariable logistic regression model, with treatment group (DOAC-switched vs DOAC-continued) as the dependent variable and the covariates described in the Baseline Covariates section as independent variables. Balance of covariates was assessed using standardized mean differences (SMDs). An SMD less than 0.1 in considered as well balanced. The proportional hazards assumption was evaluated using log–log survival plots of the weighted Kaplan–Meier curves, and no major violations were observed. Adjusted hazard ratios (aHRs) and corresponding 95% confidence intervals of each outcome of interest comparing DOAC-switch group to DOAC-continued group were estimated using weighted Cox proportional hazards models with robust sandwich type variance estimation in an intention-to-treat approach. All analysis was performed using SAS, version 9.4 (SAS Institute) and RStudio version 2025.09.0 + 418.

### Secondary Analyses

We performed 3 planned secondary analyses.

First, we compared clinical outcomes between patients who continued DOAC therapy within the same mechanism of action and those who switched to a DOAC with a different mechanism of action. Continuation within the same mechanism of action was defined as continued use of either a factor Xa inhibitor (apixaban, rivaroxaban, edoxaban) or a direct thrombin inhibitor (dabigatran), including patients who remained on the same DOAC agent or switched to a different DOAC with the same mechanism of action. In contrast, switching to a different mechanism of action was defined as switching from a factor Xa inhibitor to a direct thrombin inhibitor or from a direct thrombin inhibitor to a factor Xa inhibitor.

Second, we compared clinical outcomes between patients with antiplatelet addition and those without antiplatelet addition after the index stroke among DOAC-continued patients who were not on antiplatelet therapy before the index stroke.

Third, for patients who were treated with rivaroxaban or apixaban before the index stroke (the most commonly prescribed DOACs), we performed DOAC-specific comparisons of continuing the same agent versus switching to an alternative DOAC (e.g., among apixaban users, comparing those switching to rivaroxaban vs continuing apixaban).

For all secondary analyses, propensity score-based overlap weighting was applied to balance covariates, and weighted Cox proportional hazards regression models with robust variance estimation were used to estimate aHRs and 95% confidence intervals.

## Results

We identified 3059 AF patients who experienced an ischemic stroke while receiving DOAC therapy between January 1, 2016, and June 30, 2022 and resumed DOAC within 90 days after ischemic stroke discharge. After applying exclusion criteria, 1175 patients remained in the final study (Figure 1). The mean (SD) age was 74.7 (12.6) years, and 53.8% were males. Before the index stroke, apixaban was the most commonly prescribed DOAC (61.0%), followed by rivaroxaban (32.7%), dabigatran (6.1%), and edoxaban (0.2%). After the index stroke, apixaban continued to be the most frequently prescribed DOAC (66.9%), followed by rivaroxaban (26.6%), dabigatran (6.3%), and edoxaban (0.2%) (Figure 2). Different DOAC strategies after the index stroke were presented in Supplementary Materials (Table S2). Overall, 970 patients (82.6%) were in DOAC-continued group, whereas 205 patients (17.4%) were in DOAC-switched group.

**Figure 1.**
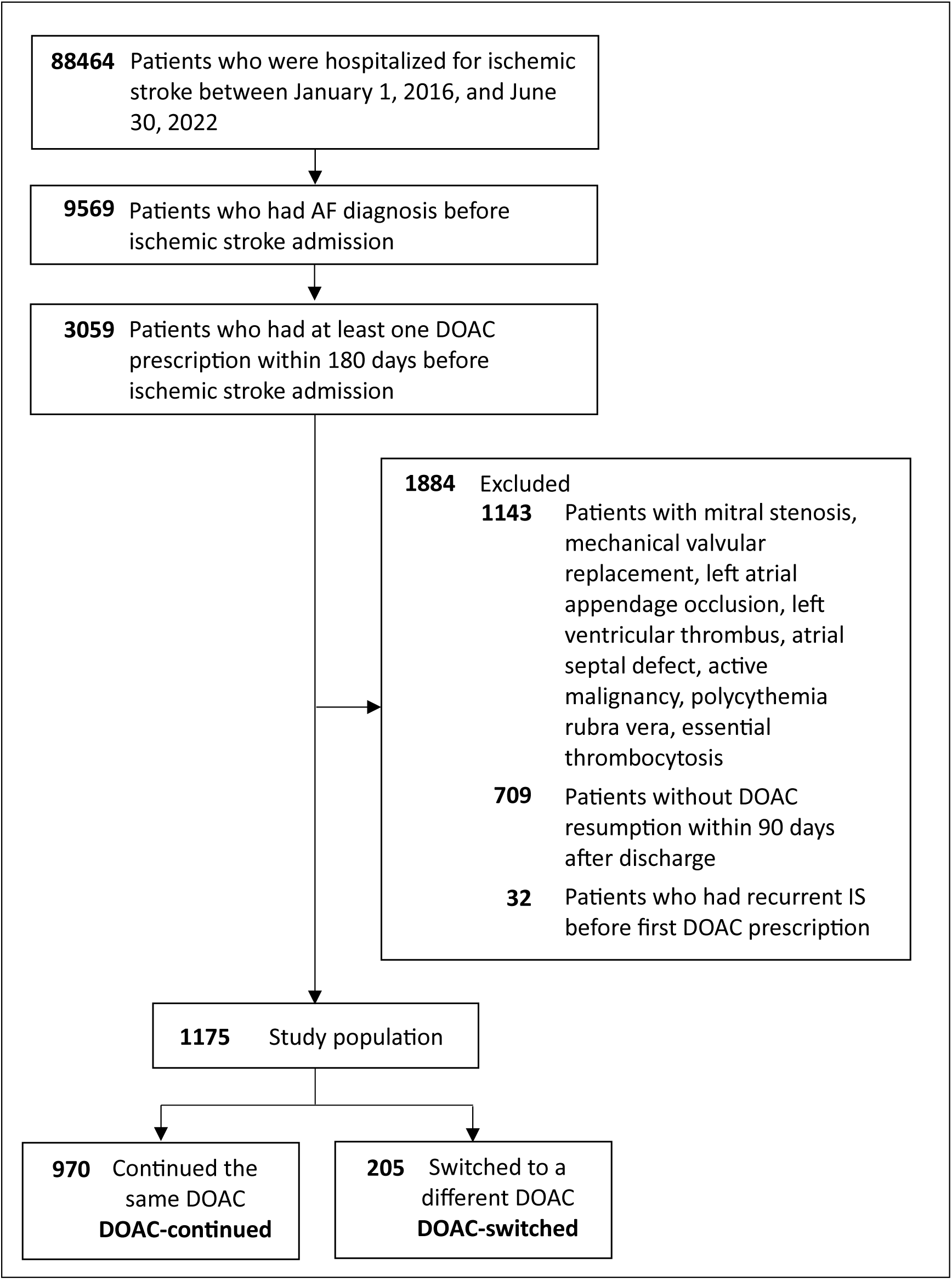
Study Population Selection Process. Abbreviation: DOAC, direct oral anticoagulant.

**Figure 2.**
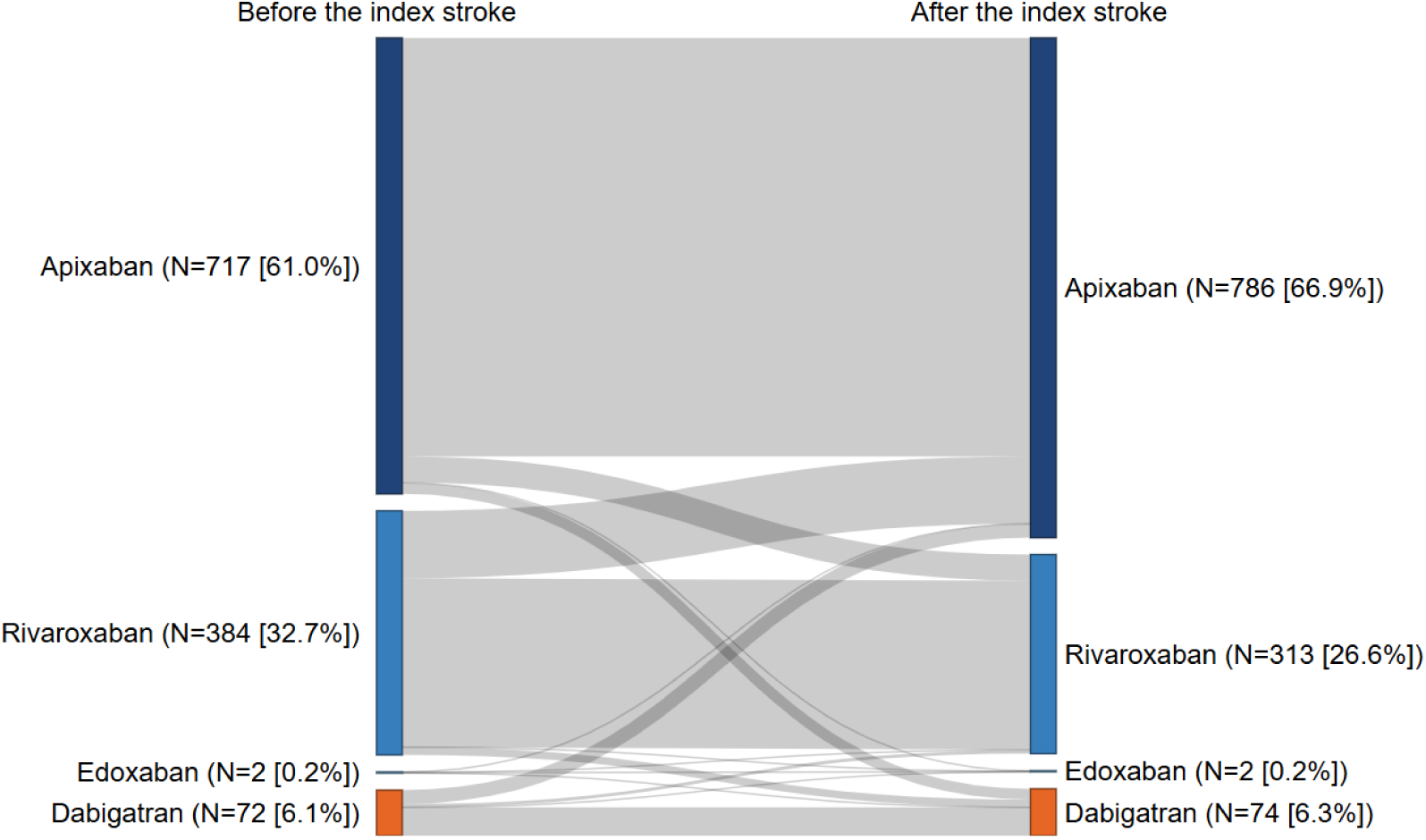
Sankey Diagram of DOAC Strategies Before and After Ischemic Stroke.

Table 1 summarizes baseline demographics, comorbidities, and medication used in the DOAC-continued and DOAC-switched groups before and after propensity score-based overlap weighting, along with ASDs. Before weighting, patients in the DOAC-switched group were more likely to be male, had a lower burden of comorbidities, lower CHA₂DS₂-VASc Score and HAS-BLED Score, and were less likely to receive CYP/P-gp modulators concomitantly compared with those in the DOAC-switched group. After weighting, baseline characteristics were well balanced between the two groups, with all ASDs below 0.1.

**Table 1.** Baseline Characteristics Before and After Propensity Score Based-Overlap Weighting. Abbreviations: ASD, absolute standardized difference; CYP, cytochrome P450; DOAC, direct oral anticoagulant; NSAIDs, nonsteroidal anti-inflammatory drugs; P-gp, p-glycoprotein; SD, standard deviation. * Indicates ASD ≥ 0.1. † Weights were derived using propensity score-based overlap weighting. Propensity scores were estimated from a logistic regression model including all variables listed in this table except the CHA_2_DS_2_-VASc and HAS-BLED scores.

|  | Before Weighting |  |  | After weighting † |  |  |
| --- | --- | --- | --- | --- | --- | --- |
|  | DOAC-switched<br>(N=205) | DOAC-continued<br>(N=970) | ASD | DOAC-switched<br>(N=166) | DOAC-continued<br>(N=166) | ASD |
| **Demographics** |  |  |  |  |  |  |
| Age, mean±SD, years | 73.7<br>(11.8) | 74.9<br>(12.8) | 0.095 | 73.9<br>(10.6) | 73.9<br>(5.5) | 0.000 |
| Male sex, n (%) | 119<br>(58.0%) | 513<br>(52.9%) | 0.104* | 95<br>(57.1%) | 95<br>(57.1%) | 0.000 |
| Drinker, n (%) | 9 (4.4) | 57 (5.9) | 0.068 | 8 (4.6) | 8 (4.6) | 0.000 |
| **Comorbidities** |  |  |  |  |  |  |
| Congestive heart failure, n (%) | 94<br>(45.9%) | 511<br>(52.7%) | 0.137* | 78<br>(47.2%) | 78<br>(47.2%) | 0.000 |
| Hypertension, n (%) | 196<br>(95.6%) | 952<br>(98.1%) | 0.146* | 160<br>(96.5%) | 160<br>(96.5%) | 0.000 |
| Diabetes mellitus, n (%) | 96<br>(46.8%) | 492<br>(50.7%) | 0.078 | 79<br>(47.4%) | 79<br>(47.4%) | 0.000 |
| Coronary artery disease, n (%) | 102<br>(49.8%) | 577<br>(59.5%) | 0.196* | 85<br>(51.4%) | 85<br>(51.4%) | 0.000 |
| Peripheral artery disease, n (%) | 44<br>(21.5%) | 195<br>(20.1%) | 0.034 | 36<br>(21.5%) | 36<br>(21.5%) | 0.000 |
| Venous thromboembolism, n (%) | 14 (6.8%) | 88 (9.1%) | 0.083 | 12 (7%) | 12 (7%) | 0.000 |
| Chronic kidney disease, n (%) | 72<br>(35.1%) | 387<br>(39.9%) | 0.099 | 60<br>(36.1%) | 60<br>(36.1%) | 0.000 |
| Chronic liver disease, n (%) | 17 (8.3%) | 73 (7.5%) | 0.028 | 13 (8%) | 13 (8%) | 0.000 |
| Anemia, n (%) | 66<br>(32.2%) | 361<br>(37.2%) | 0.106* | 54<br>(32.7%) | 54<br>(32.7%) | 0.000 |
| Hyperlipidemia, n (%) | 200<br>(97.6%) | 949<br>(97.8%) | 0.019 | 162<br>(97.6%) | 162<br>(97.6%) | 0.000 |
| CHA<sub>2</sub>DS<sub>2</sub>-VASc Score, mean±SD, points | 5.2 (1.5) | 5.5 (1.5) | 0.220* | 5.2 (1.3) | 5.2 (0.6) | 0.020 |
| HAS-BLED Score, mean±SD, points | 3.1 (1.2) | 3.3 (1.3) | 0.186* | 3.1 (1.1) | 3.1 (0.5) | 0.039 |
| **Concomitant medication use** |  |  |  |  |  |  |
| Statin, n (%) | 185<br>(90.2%) | 848<br>(87.4%) | 0.09 | 149<br>(89.8%) | 149<br>(89.8%) | 0.000 |
| NSAIDs, n (%) | 22<br>(10.7%) | 94 (9.7%) | 0.034 | 17<br>(10.5%) | 17<br>(10.5%) | 0.000 |
| Proton pump inhibitors, n (%) | 54<br>(26.3%) | 263<br>(27.1%) | 0.017 | 44<br>(26.4%) | 44<br>(26.4%) | 0.000 |
| CYP/P-gp modulators, n (%) | 15 (7.3%) | 111<br>(11.4%) | 0.142* | 13<br>(7.9%) | 13 (7.9%) | 0.000 |
| Antiplatelet before first ischemic stroke, n (%) | 15 (7.3%) | 81 (8.4%) | 0.038 | 13<br>(7.6%) | 13 (7.6%) | 0.000 |
| Antiplatelet after first ischemic stroke, n (%) | 31<br>(15.1%) | 136<br>(14%) | 0.031 | 25<br>(14.8%) | 25<br>(14.8%) | 0.000 |

The median follow-up was 10.2 months in the DOAC-switched group and 8.9 months in the DOAC-continued group. Recurrent ischemic stroke occurred in 12 of 205 and 44 of 970 patients, corresponding to incidence rates of 3.9 and 3.4 per 1000 person-months, respectively (Table 2). For DOAC-switched group versus DOAC-continued group, there was no significant difference in the hazard of recurrent ischemic stroke (aHR, 1.20; 95% CI, 0.63-2.30), major bleeding (aHR, 0.60; 95% CI, 0.21-1.72), and composite outcome (aHR, 0.98; 95% CI, 0.56-1.70) (Figure 3, S1-S3).

**Figure 3.**
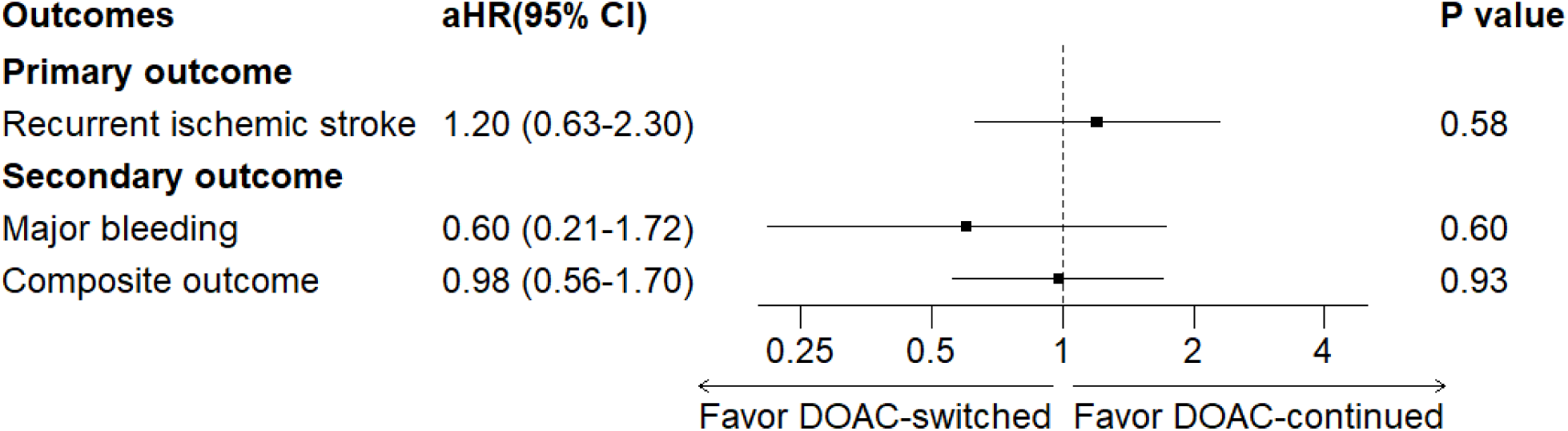
Adjusted Hazard Ratios for Clinical Outcomes Comparing Switching to a Different DOAC (DOAC-switched, N=205) with Continuing the Same DOAC (DOAC-continued, N=970) After Ischemic Stroke. aHRs are plotted on a logarithmic scale. Abbreviations: DOAC, direct oral anticoagulant; aHR, adjusted hazard ratio; CI, confidence interval

**Table 2.** Clinical Outcomes Comparing DOAC-switched (N=205) vs. DOAC-continued (N=970) Abbreviation: DOAC, direct oral anticoagulant. * Weighted incidence rates were calculated as the sum of weighted event counts divided by the sum of weighted person-months.

| Outcome | Exposure | Number of events | Person-months | Median follow-up month | Incidence rate (per 1000 person-months) |  |
| --- | --- | --- | --- | --- | --- | --- |
|  |  |  |  |  | Unweighted | Weighted * |
| Recurrent ischemic stroke | DOAC-switched | 12 | 3024.3 | 10.2 | 4.0 | 3.9 |
|  | DOAC-continued | 44 | 12644.7 | 8.9 | 3.5 | 3.4 |
| Major bleeding | DOAC-switched | 4 | 3119.4 | 11.5 | 1.3 | 1.3 |
|  | DOAC-continued | 30 | 12732.1 | 9.2 | 2.4 | 2.2 |
| Composite outcome | DOAC-switched | 16 | 3005.1 | 10.2 | 5.3 | 5.3 |
|  | DOAC-continued | 73 | 12410.8 | 8.6 | 5.9 | 5.6 |

### Secondary Analyses

Among the 1175 patients, 1119 patients (95.2%) continued a DOAC with the same mechanism of action, whereas 56 patients (4.7%) switched to a DOAC with a different mechanism of action. Compared with continuing a DOAC with the same mechanism of action, switching to a DOAC with a different mechanism of action was not significantly associated with recurrent ischemic stroke (aHR, 1.43; 95% CI, 0.50-4.11), major bleeding (aHR, 0.62; 95% CI, 0.08-4.82), or composite outcome (aHR, 1.16; 95% CI, 0.46-2.97) (Figure 4, Table S3, Figure S1).

**Figure 4.**
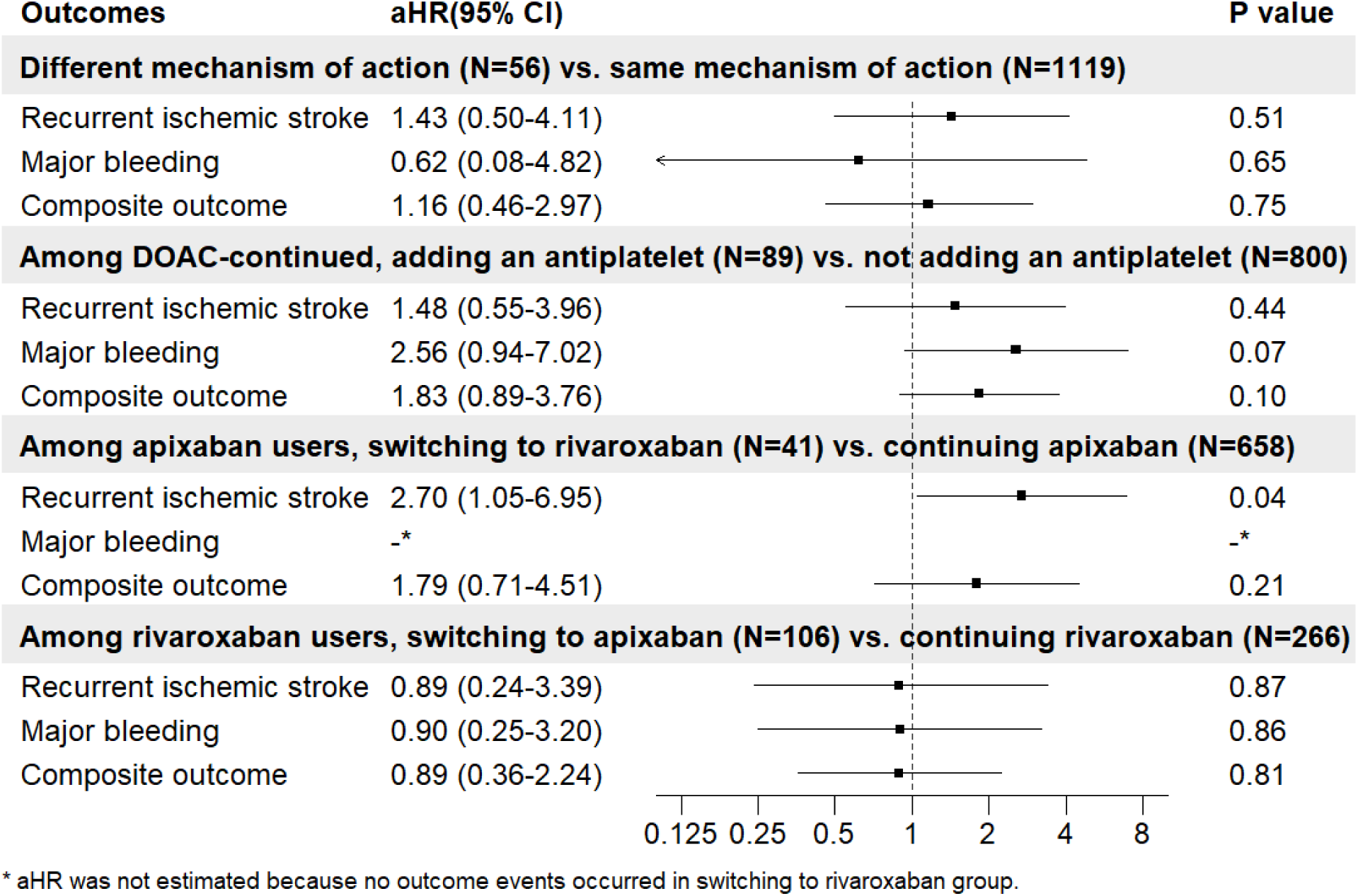
Adjusted Hazard Ratios for Clinical Outcomes Comparing Different DOAC Strategies. aHRs are plotted on a logarithmic scale. Abbreviations: DOAC, direct oral anticoagulant; aHR, adjusted hazard ratio; CI, confidence interval

Of the 889 patients in the DOAC-continued group who were not on antiplatelet therapy prior to the index stroke, 89 (10.0%) initiated antiplatelet therapy after the index stroke, while 800 (90.0%) remained without antiplatelet therapy. Compared with patients without antiplatelet addition, those with antiplatelet addition did not have a significantly lower risk of recurrent ischemic stroke (aHR, 1.48; 95% CI, 0.55-3.96), a significantly higher risk of major bleeding (aHR, 2.56; 95% CI, 0.94-7.02), or a significantly different risk of the composite outcome (aHR, 1.83; 95% CI, 0.89-3.76) (Figure 4, Table S4).

Among 717 patients who received apixaban before the index stroke, 658 (91.8%) continued apixaban, 41 (5.7%) switched to rivaroxaban, 1 (0.1%) switched to edoxaban, and 17 (2.4%) switched to dabigatran (Table S2). Compared with continuing apixaban, switching to rivaroxaban was associated with a higher hazard of recurrent ischemic stroke (aHR, 2.70; 95% CI, 1.05–6.95) (Figure 4, Table S5). The hazard ratio for bleeding could not be estimated because no bleeding events occurred among patients who switched to rivaroxaban. Comparisons for switching from apixaban to edoxaban or dabigatran were not performed because of limited sample size.

Among 384 patients who received rivaroxaban before the index stroke, 266 (69.3%) continued rivaroxaban, 106 (27.6%) switched to apixaban, none switched to edoxaban, and 12 (3.1%) switched to dabigatran (Table S2). Compared with continuing rivaroxaban, switching to apixaban was not associated with a significantly different risk of recurrent ischemic stroke (aHR, 0.89; 95% CI, 0.24–3.39), major bleeding (aHR, 0.90; 95% CI, 0.25– 3.20), or the composite outcome (aHR, 0.89; 95% CI, 0.36–2.24) (Figure 4, Table S6). Comparisons for switching from rivaroxaban to edoxaban or dabigatran were not performed because of limited sample size.

Analyses among patients who received edoxaban or dabigatran before the index stroke were not conducted because of small sample sizes and limited outcome events.

## Discussion

In this study, we investigated the effect of different DOAC strategies on clinical outcomes among patients who experienced ischemic stroke while on DOAC therapy and resumed DOAC within 90 days after ischemic stroke discharge in the United States using administrative claim data. We found that neither switching to a different DOAC (vs. continuing the same DOAC) nor switching to a DOAC with a different mechanism of action (vs. continuing the same mechanism) was associated with recurrent ischemic stroke or major bleeding events. When restricting our analyses to patients who continued the same DOAC and did not receive antiplatelet before the index stroke, we found that adding antiplatelet after the index stroke was not associated with a lower risk of recurrent ischemic stroke. Among patients who received apixaban before the index stroke, switching to rivaroxaban (vs. continuing apixaban) after the index stroke was associated with a higher risk of recurrent ischemic stroke. Within patients who received rivaroxaban before the index stroke, switching to apixaban (vs. continuing rivaroxaban) after the index stroke was associated with slightly lower risk of recurrent ischemic stroke, although the difference was not statistically significant.

Recent meta-analyses of observational cohort studies conducted across multiple countries have reported that the risks of recurrent ischemic stroke, intracranial hemorrhage, and major bleeding were similar between patients who switched to another DOAC and those who continued the same DOAC. ^18,19^ Furthermore, initiation of antiplatelet therapy after the index stroke did not reduce the risk of recurrent ischemic stroke. ^18^ Switching to a DOAC with a different mechanism of action was also not associated with a lower risk of recurrent ischemic stroke or intracranial hemorrhage.^18^ Our findings are consistent with these meta-analyses and further support current guideline recommendations, which do not suggest adding antiplatelet therapy or switching anticoagulants in patients who experience an ischemic stroke while on anticoagulation therapy. ^7^

Similar differences among individual DOACs have been reported previously. A meta-analysis of observational studies comparing apixaban and rivaroxaban in AF patients found lower risks of both stroke or systemic embolism and major bleeding with apixaban, with a greater relative benefit for thromboembolic outcomes observed in studies with higher mean CHA₂DS₂-VASc scores. ^20^ Notably, the studies included in that meta-analysis comprised a mixture of patients with and without prior ischemic stroke and therefore reflected both primary and secondary stroke prevention settings. Although no head-to-head randomized trial comparing apixaban and rivaroxaban has been completed in patients with atrial fibrillation, recent randomized evidence in patients with acute venous thromboembolism similarly demonstrated lower rates of clinically relevant bleeding with apixaban than with rivaroxaban.^21^ Despite the differences in study populations, both studies suggest that clinically meaningful differences may exist among individual DOACs. Consistent with these observations, we found that switching from apixaban to rivaroxaban was associated with a significantly higher risk of recurrent ischemic stroke, whereas switching from rivaroxaban to apixaban was associated with slightly lower risks of recurrent ischemic stroke and bleeding, although statistical significance was not reached. Taken together, these findings suggest that outcomes may be influenced less by whether a DOAC is switched and more by which DOAC is selected after the stroke. Future studies with larger sample sizes and longer follow-up durations, particularly randomized controlled trials, are warranted to compare individual DOAC agents for secondary stroke prevention in AF patients.

One strength of this study is the use of real-world data from administrative claims including exclusively United States population. In addition to evaluation the outcomes of switching versus continuing same type of DOAC therapy or DOAC with same mechanism of action after ischemic stroke, we further performed exploratory analyses comparing individual DOAC agents. This approach provides an additional framework for understanding post-stroke anticoagulation strategies and allows assessment of whether clinical outcomes may differ among specific DOACs.

Another strength is the use of overlap weighting to address baseline differences in treatment selection. Because dosing adjustment criteria, renal function thresholds, and other prescribing considerations vary across individual DOACs, not all patients are equally eligible to receive every available agent. Recent observational studies have shown that apixaban users tend to be older and have a greater burden of comorbidities, as well as higher CHA₂DS₂-VASc and HAS-BLED scores ^22^, suggesting systematic differences in treatment allocation across DOACs. By applying overlap weighting, we estimated the average treatment effect in the overlap population (ATO), focusing on patients for whom multiple DOAC options were clinically plausible and treatment assignment was less strongly influenced by contraindications or extreme clinical characteristics. Consequently, our findings are most applicable to patients within this overlap population and should not be interpreted as representing the treatment effect in all patients with atrial fibrillation receiving DOAC therapy.

However, several limitations should be considered. The MarketScan database includes insured employees, their dependents, early retirees, COBRA continuers, and Medicare-eligible retirees, but does not capture Medicaid enrollees or uninsured individuals, which may limit the generalizability of our findings. In addition, the database lacks information on medication adherence, body weight, and laboratory measurements, including renal function, precluding assessment of treatment adherence and the appropriateness of DOAC dosing. Stroke mechanisms could not be reliably determined from claims data. The coexistence of diagnoses of atrial fibrillation and ischemic stroke does not necessarily indicate a cardioembolic etiology, and competing stroke mechanisms may have contributed to either the index or recurrent ischemic stroke events. Consequently, we were unable to determine whether ischemic stroke events occurring during DOAC therapy were attributable to atrial fibrillation–related cardioembolism, competing stroke mechanisms, inadequate adherence, or inappropriate dosing. Nevertheless, we emphasize that in clinical practice, these factors should be carefully evaluated before considering making changes to DOAC therapy, in accordance with current guideline recommendations. In addition, number of endpoints was limited for most comparisons, resulting in imprecise estimates of association. Finally, the database does not contain information on race, ethnicity, socioeconomic status, or mortality, raising the possibility of residual confounding and preventing assessment of mortality outcomes.

In conclusion, switching DOAC therapy after ischemic stroke was not associated with improved clinical outcomes. Switching from apixaban to rivaroxaban, however, could increase risk of recurrent ischemic stroke. Further studies are needed to directly compare individual DOAC agents in secondary stroke.

## Data Availability

Due to licensing restrictions, all data produced in the present study cannot be made available to other investigators to reproduce results, but researchers may contact Merative to obtain and license the data.

## Acknowledgments

None

## Supplemental Material

Table S1–S6

Figure S1–S5

## Non-standard Abbreviations and Acronyms

DOAC: direct oral anticoagulant
AF: atrial fibrillation

