## Supplementary material for "Clinical Outcomes of Switching vs. Continuing Direct Oral Anticoagulants (DOACs) After Ischemic Stroke in Patients with Atrial Fibrillation in the US": Table S1-S6, Figure S1-S5

### Supplementary Materials

Supplementary Table 1. Diagnosis Code and Procedure Code for Inclusion Criteria, Exclusion Criteria, Outcomes, and Covariates.

* Using definition from Chronic Conditions Data Warehouse from the Center for Medicare and Medicaid Services (https://www2.ccwdata.org/web/guest/condition-categories-chronic)

Abbreviations: CPT-4, Current Procedural Terminology, Fourth Edition; DOAC, direct oral anticoagulant; ICD-10-CM, International Classification of Diseases, Tenth Revision, Clinical Modification; ICD-10-PCS, International Classification of Diseases, Tenth Revision, Procedure Coding System.

| Diagnosis/procedure | ICD-10-CM/ICD-10-PCS/CPT-4 |
|---|---|
| Inclusion criteria | |
| Ischemic stroke | I63 |
| Atrial fibrillation | I48 |
| Exclusion criteria | |
| Mitral stenosis | I34.2, I05.0, I05.2, Q23.2 |
| Mechanical valve replacement | Z95.2 |
| Left ventricular thrombus | I51.3 |
| Atrial septal defect | Q21.1<br>Q21.2 |
| Active malignancy | Code that starts with “C” |
| Polycythemia vera | D45 |
| Essential thrombocytosis | D47.3 |
| Percutaneous/surgical left atrial appendage occlusion | ICD-10-PCS: 02L70CK, 02L70DK, 02L70ZK, 02L73CK, 02L73DK, 02L73ZK, 02L74CK, 02L74DK, 02L74ZK<br>CPT-4: 33340 |
| Primary outcome | |
| Ischemic stroke | I63 |
| Secondary outcome | |
| Intracranial hemorrhage | I60, I61, I62 |
| Gastrointestinal bleeding | I85.01, I85.11, K20.81, K20.91, K22.11, K22.6, K25.0, K25.2, K25.4, K25.6, K26.0, K26.2, K26.4, K26.6, K27.0, K27.2, K27.4, K27.6, K28.0, K28.2, K28.4, K28.6, K29.01, K29.21, K29.31, K29.41, K29.51, K29.61, K29.71, K29.81, K29.91, K31.811, K31.82, |

| | |
|---|---|
| | K55.21, K57.01, K57.11, K57.13, K57.21, K57.31, K57.33, K57.41, K57.51, K57.53, K57.81, K57.91, K57.93, K62.5, K92.0, K92.1, K92.2 |
| Other bleeding | D69.8, D69.9, I31.2, M25.0, R04, R31.0, R31.9, R58 |
| Covariates | |
| Items in CHA2DS2-VASc score | |
| Congestive heart failure | I09.81, I11.0, I13.0, I13.2, I50.1, I50.20, I50.21, I50.22, I50.23, I50.30, I50.31, I50.32, I50.33, I50.40, I50.41, I50.42, I50.43, I50.810, I50.811, I50.812, I50.813, I50.814, I50.82, I50.83, I50.84, I50.89, I50.9, P29.0 |
| Hypertension | H35.031, H35.032, H35.033, H35.039, I10, I11.0, I11.9, I12.0, I12.9, I13.0, I13.10, I13.11, I13.2, I15.0, I15.1, I15.2, I15.8, I15.9, I1A.0, I67.4, N26.2 |
| Diabetes * | E08.00, E08.01, E08.10, E08.11, E08.21, E08.22, E08.29, E08.311, E08.319, E08.321, E08.3211, E08.3212, E08.3213, E08.3219, E08.329, E08.3291, E08.3292, E08.3293, E08.3299, E08.331, E08.3311, E08.3312, E08.3313, E08.3319, E08.339, E08.3391, E08.3392, E08.3393, E08.3399, E08.341, E08.3411, E08.3412, E08.3413, E08.3419, E08.349, E08.3491, E08.3492, E08.3493, E08.3499, E08.351, E08.3511, E08.3512, E08.3513, E08.3519, E08.3521, E08.3522, E08.3523, E08.3529, E08.3531, E08.3532, E08.3533, E08.3539, E08.3541, E08.3542, E08.3543, E08.3549, E08.3551, E08.3552, E08.3553, E08.3559, E08.359, E08.3591, E08.3592, E08.3593, E08.3599, E08.36, E08.37X1, E08.37X2, E08.37X3, E08.37X9, E08.39, E08.40, E08.41, E08.42, E08.43, E08.44, E08.49, E08.51, E08.52, E08.59, E08.610, E08.618, E08.620, E08.621, E08.622, E08.628, E08.630, E08.638, E08.641, E08.649, E08.65, E08.69, E08.8, E08.9, E09.00, E09.01, E09.10, E09.11, E09.21, E09.22, E09.29, E09.311, E09.319, E09.321, E09.3211, E09.3212, E09.3213, E09.3219, E09.329, E09.3291, E09.3292, E09.3293, E09.3299, E09.331, E09.3311, E09.3312, E09.3313, E09.3319, E09.339, E09.3391, E09.3392, E09.3393, E09.3399, E09.341, E09.3411, E09.3412, E09.3413, E09.3419, E09.349, E09.3491, E09.3492, E09.3493, E09.3499, E09.351, E09.3511, E09.3512, E09.3513, E09.3519, E09.3521, E09.3522, E09.3523, E09.3529, E09.3531, E09.3532, E09.3533, E09.3539, E09.3541, E09.3542, E09.3543, E09.3549, E09.3551, E09.3552, |

| | |
|---|---|
| | E09.3553, E09.3559, E09.359, E09.3591, E09.3592, E09.3593, E09.3599, E09.36, E09.37X1, E09.37X2, E09.37X3, E09.37X9, E09.39, E09.40, E09.41, E09.42, E09.43, E09.44, E09.49, E09.51, E09.52, E09.59, E09.610, E09.618, E09.620, E09.621, E09.622, E09.628, E09.630, E09.638, E09.641, E09.649, E09.65, E09.69, E09.8, E09.9, E10.10, E10.11, E10.21, E10.22, E10.29, E10.311, E10.319, E10.321, E10.3211, E10.3212, E10.3213, E10.3219, E10.329, E10.3291, E10.3292, E10.3293, E10.3299, E10.331, E10.3311, E10.3312, E10.3313, E10.3319, E10.339, E10.3391, E10.3392, E10.3393, E10.3399, E10.341, E10.3411, E10.3412, E10.3413, E10.3419, E10.349, E10.3491, E10.3492, E10.3493, E10.3499, E10.351, E10.3511, E10.3512, E10.3513, E10.3519, E10.3521, E10.3522, E10.3523, E10.3529, E10.3531, E10.3532, E10.3533, E10.3539, E10.3541, E10.3542, E10.3543, E10.3549, E10.3551, E10.3552, E10.3553, E10.3559, E10.359, E10.3591, E10.3592, E10.3593, E10.3599, E10.36, E10.37X1, E10.37X2, E10.37X3, E10.37X9, E10.39, E10.40, E10.41, E10.42, E10.43, E10.44, E10.49, E10.51, E10.52, E10.59, E10.610, E10.618, E10.620, E10.621, E10.622, E10.628, E10.630, E10.638, E10.641, E10.649, E10.65, E10.69, E10.8, E10.9, E10.A0, E10.A1, E10.A2 |
| Coronary artery disease | I20, I21, I22, I25.1, I25.2, Z95.1, Z95.5 |
| Peripheral artery disease | I70, I74 |
| Venous thromboembolism | I82 |
| Items in HAS-BLED score | |
| Chronic kidney disease * | A18.11, A52.75, B52.0, E08.21, E08.22, E08.29, E09.21, E09.22, E09.29, E10.21, E10.22, E10.29, E11.21, E11.22, E11.29, E13.21, E13.22, E13.29, I12.0, I12.9, I13.0, I13.10, I13.11, I13.2, K76.7, M10.30, M10.311, M10.312, M10.319, M10.321, M10.322, M10.329, M10.331, M10.332, M10.339, M10.341, M10.342, M10.349, M10.351, M10.352, M10.359, M10.361, M10.362, M10.369, M10.371, M10.372, M10.379, M10.38, M10.39, M32.14, M32.15, M35.04, M35.0A, N01.0, N01.1, N01.2, N01.3, N01.4, N01.5, N01.6, N01.7, N01.8, N01.9, N01.A, N02.0, N02.1, N02.2, N02.3, N02.4, N02.5, N02.6, N02.7, N02.8, N02.9, N02.A, N02.B1, N02.B2, N02.B3, N02.B4, N02.B5, N02.B6, N02.B9, N03.0, N03.1, N03.2, N03.3, N03.4, N03.5, N03.6, N03.7, N03.8, N03.9, N03.A, N04.0, N04.1, N04.2, N04.20, N04.21, N04.22, N04.29, N04.3, |

| | |
|---|---|
| | N04.4, N04.5, N04.6, N04.7, N04.8, N04.9, N04.A, N05.0, N05.1, N05.2, N05.3, N05.4, N05.5, N05.6, N05.7, N05.8, N05.9, N05.A, N06.0, N06.1, N06.2, N06.20, N06.21, N06.22, N06.29, N06.3, N06.4, N06.5, N06.6, N06.7, N06.8, N06.9, N06.A, N07.0, N07.1, N07.2, N07.3, N07.4, N07.5, N07.6, N07.7, N07.8, N07.9, N07.A, N08, N14.0, N14.1, N14.11, N14.19, N14.2, N14.3, N14.4, N15.0, N15.8, N15.9, N16, N18.1, N18.2, N18.3, N18.30, N18.31, N18.32, N18.4, N18.5, N18.6, N18.9, N25.1, N25.89, N25.9, N26.1, N26.9, N99.0, Q61.02, Q61.11, Q61.19, Q61.2, Q61.3, Q61.4, Q61.5, Q61.8, Y84.1, Z49.01, Z49.02, Z49.31, Z49.32 |
| Chronic liver disease | K70, K71, K72, K73, K74, K75, K76, K77 |
| Anemia * | C94.6, D46.0, D46.1, D46.20, D46.21, D46.22, D46.4, D46.9, D46.A, D46.B, D46.C, D46.Z, D47.4, D50.0, D50.1, D50.8, D50.9, D51.0, D51.1, D51.2, D51.3, D51.8, D51.9, D52.0, D52.1, D52.8, D52.9, D53.0, D53.1, D53.2, D53.8, D53.9, D55.0, D55.1, D55.2, D55.21, D55.29, D55.3, D55.8, D55.9, D56.0, D56.1, D56.2, D56.3, D56.4, D56.5, D56.8, D56.9, D57.00, D57.01, D57.02, D57.03, D57.04, D57.09, D57.1, D57.20, D57.211, D57.212, D57.213, D57.214, D57.218, D57.219, D57.3, D57.40, D57.411, D57.412, D57.413, D57.414, D57.418, D57.419, D57.42, D57.431, D57.432, D57.433, D57.434, D57.438, D57.439, D57.44, D57.451, D57.452, D57.453, D57.454, D57.458, D57.459, D57.80, D57.811, D57.812, D57.813, D57.814, D57.818, D57.819, D58.0, D58.1, D58.2, D58.8, D58.9, D59.0, D59.1, D59.10, D59.11, D59.12, D59.13, D59.19, D59.2, D59.3, D59.30, D59.31, D59.32, D59.39, D59.4, D59.5, D59.6, D59.8, D59.9, D60.0, D60.1, D60.8, D60.9, D61.01, D61.02, D61.09, D61.1, D61.2, D61.3, D61.810, D61.811, D61.818, D61.82, D61.89, D61.9, D63.0, D63.1, D63.8, D64.0, D64.1, D64.2, D64.3, D64.4, D64.81, D64.89, D64.9, D75.81 |
| Alcohol use (drinker) | F10, G62.1, I42.6, K29.2, K70, T51.0, T51.9, Z71.4 |
| Others | |
| Hyperlipidemia * | E78.0, E78.00, E78.01, E78.1, E78.2, E78.3, E78.4, E78.41, E78.49, E78.5 |

Table S2. Different DOAC Treatment Strategies

Abbreviations: DOAC, direct oral anticoagulant.

| Patients | Strategies | | N (%) |
|---|---|---|---|
| All patients (N=1175) | Before the index stroke | After the index stroke | |
| | Apixaban | Apixaban | 658 (56.0%) |
| | Apixaban | Rivaroxaban | 41 (3.5%) |
| | Apixaban | Edoxaban | 1 (0.1%) |
| | Apixaban | Dabigatran | 17 (1.5%) |
| | Rivaroxaban | Apixaban | 106 (9.0%) |
| | Rivaroxaban | Rivaroxaban | 266 (22.6%) |
| | Rivaroxaban | Edoxaban | 0 (0.0%) |
| | Rivaroxaban | Dabigatran | 12 (1.0%) |
| | Edoxaban | Apixaban | 0 (0.0%) |
| | Edoxaban | Rivaroxaban | 1 (0.1%) |
| | Edoxaban | Edoxaban | 1 (0.1%) |
| | Edoxaban | Dabigatran | 0 (0.0%) |
| | Dabigatran | Apixaban | 22 (1.9%) |
| | Dabigatran | Rivaroxaban | 5 (0.4%) |
| | Dabigatran | Edoxaban | 0 (0.0%) |
| | Dabigatran | Dabigatran | 45 (3.8%) |
| All patients (N=1175) | Continued the same type of DOAC after the index stroke (DOAC-continued) | | 970 (82.6%) |
| | Switched to a different type of DOAC after the index stroke (DOAC-switched) | | 205 (17.4%) |
| All patients (N=1175) | Continued a DOAC with the same mechanism of action after the index stroke | | 1119 (95.2%) |
| | Switched to a DOAC with a different mechanism of action after the index stroke | | 56 (4.7%) |
| DOAC-continued patients who were not on antiplatelet therapy before the index stroke (N=889) | Without antiplatelet addition after index the index stroke | | 800 (90.0%) |
| | With antiplatelet addition after index the index stroke | | 89 (10.0%) |

| | | |
|---|---|---|
| Patients who received apixaban before the index stroke(N=717) | After the index stroke | |
| | Apixaban | 658 (91.8%) |
| | Rivaroxaban | 41 (5.7%) |
| | Edoxaban | 1 (0.1%) |
| | Dabigatran | 17 (2.4%) |
| Patients who received rivaroxaban before the index stroke(N=384) | After the index stroke | |
| | Apixaban | 106(27.6%) |
| | Rivaroxaban | 266(69.3%) |
| | Edoxaban | 0 (0.0%) |
| | Dabigatran | 12 (3.1%) |
| Patients who received edoxaban before the index stroke(N=2) | After the index stroke | |
| | Apixaban | 0 (0.0%) |
| | Rivaroxaban | 1 (50.0%) |
| | Edoxaban | 1 (50.0%) |
| | Dabigatran | 0 (0.0%) |
| Patients who received dabigatran before the index stroke(N=72) | After the index stroke | |
| | Apixaban | 22 (30.6%) |
| | Rivaroxaban | 5 (6.9%) |
| | Edoxaban | 0 (0.0%) |
| | Dabigatran | 45 (62.5%) |

Table S3. Clinical Outcomes Comparing Switching to a DOAC with a Different Mechanism of Action(N=56) with Continuing a DOAC with the Same Mechanism of Action (N=1119) After Ischemic Stroke

Abbreviation: aHR, adjusted hazard ratio.

* Weighted incidence rates were calculated as the sum of weighted event counts divided by the sum of weighted person-months.

† Adjusted hazard ratios (aHRs) were estimated using Cox proportional hazards regression with propensity score–based overlap weighting. The overlap weights were derived from a propensity score model accounting for age, sex, alcohol drinker, congestive heart failure, hypertension, diabetes mellitus, coronary artery disease, peripheral artery disease, venous thromboembolism, chronic liver disease, anemia, hyperlipidemia, concomitant statin, NSAID, proton pump inhibitor, CYP/P-gp modulator use, and antiplatelet use before and after the index stroke.

| Outcome | Exposure | Number of events | Person-months | Median follow-up month | Incidence rate (per 1000 person-months) | | aHR (95% CI) † | P value |
|---|---|---|---|---|---|---|---|---|
| | | | | | unweighted | Weighted * | | |
| Recurrent ischemic stroke | Different mechanism | 4 | 1017.3 | 11.9 | 3.9 | 3.9 | 1.43 (0.50-4.11) | 0.51 |
| | Same mechanism | 52 | 14651.7 | 9.0 | 3.5 | 3.2 | (reference) | |
| Major bleeding | Different mechanism | 1 | 1026.6 | 13.0 | 1.0 | 1.0 | 0.62 (0.08-4.82) | 0.65 |
| | Same mechanism | 33 | 14824.8 | 9.3 | 2.2 | 1.8 | (reference) | |
| Composite outcome | Different mechanism | 5 | 1010.9 | 11.9 | 4.9 | 4.9 | 1.16 (0.46 - 2.97) | 0.75 |
| | Same mechanism | 84 | 14405.1 | 8.83778 | 5.8 | 5.1 | (reference) | |

Table S4. Clinical Outcomes Comparing Addition of an Antiplatelet (N=89) with No Antiplatelet Addition (N=800) Among Patients Who Continued the Same DOAC After Ischemic Stroke and Had No Prior Antiplatelet Therapy (N=889)

Abbreviation: aHR, adjusted hazard ratio.

* Weighted incidence rates were calculated as the sum of weighted event counts divided by the sum of weighted person-months.

† Adjusted hazard ratios (aHRs) were estimated using Cox proportional hazards regression with propensity score–based overlap weighting. The overlap weights were derived from a propensity score model accounting for age, sex, alcohol drinker, congestive heart failure, hypertension, diabetes mellitus, coronary artery disease, peripheral artery disease, venous thromboembolism, chronic liver disease, anemia, hyperlipidemia, concomitant statin, NSAID, proton pump inhibitor, CYP/P-gp modulator use.

| Outcome | Exposure | Number of events | Person-months | Median follow-up month | Incidence rate (per 1000 person-months) | | aHR (95% CI) † | P value |
|---|---|---|---|---|---|---|---|---|
| | | | | | unweighted | Weighted * | | |
| Recurrent ischemic stroke | With antiplatelet addition | 6 | 1187.0 | 11.9 | 5.1 | 4.7 | 1.48 (0.55-3.96) | 0.44 |
| | Without antiplatelet addition | 34 | 10620.3 | 8.8 | 3.2 | 3.3 | (reference) | |
| Major bleeding | With antiplatelet addition | 6 | 1176.1 | 12.8 | 5.1 | 6.0 | 2.56 (0.94-7.02) | 0.07 |
| | Without antiplatelet addition | 24 | 10664.7 | 9.1 | 2.3 | 2.2 | (reference) | |
| Composite outcome | With antiplatelet addition | 11 | 1131.7 | 10.8 | 9.7 | 10.6 | 1.83 (0.89-3.76) | 0.10 |
| | Without antiplatelet addition | 58 | 10441.7 | 8.5 | 5.6 | 5.6 | (reference) | |

Table S5. Clinical Outcomes Comparing Switching to Rivaroxaban (N=41) with Continuing Apixaban (N=658) Among Patients Who Received Apixaban Before Ischemic Stroke

Abbreviation: aHR, adjusted hazard ratio.

* Weighted incidence rates were calculated as the sum of weighted event counts divided by the sum of weighted person-months.

† Adjusted hazard ratios (aHRs) were estimated using Cox proportional hazards regression with propensity score–based overlap weighting. The overlap weights were derived from a propensity score model accounting for age, sex, alcohol drinker, congestive heart failure, hypertension, diabetes mellitus, coronary artery disease, peripheral artery disease, venous thromboembolism, chronic liver disease, anemia, hyperlipidemia, concomitant statin, NSAID, proton pump inhibitor, CYP/P-gp modulator use, and antiplatelet use before and after the index stroke.

‡ HR was not estimated because no outcome events occurred in switching to rivaroxaban group.

| Outcome | Exposure | Number of events | Person-months | Median follow-up month | Incidence rate (per 1000 person-months) | | aHR (95% CI) † | P value |
|---|---|---|---|---|---|---|---|---|
| | | | | | unweighted | Weighted * | | |
| Recurrent ischemic stroke | Switched to rivaroxaban | 5 | 396.9 | 7.3 | 12.6 | 12.0 | 2.70 (1.05-6.95) | 0.04 |
| | Continued apixaban | 35 | 7854.0 | 8.1 | 4.5 | 3.7 | (reference) | |
| Major bleeding | Switched to rivaroxaban | 0 | 441.0 | 9.26489 | 0 | 0 | - ‡ | - ‡ |
| | Continued apixaban | 20 | 7954.9 | 8.16427 | 2.5 | 2.1 | (reference) | |
| Composite outcome | Switched to rivaroxaban | 5 | 396.9 | 7.260780 | 12.6 | 12.0 | 1.79 (0.71-4.51) | 0.21 |
| | Continued apixaban | 54 | 7703.6 | 7.91786 | 7.0 | 5.7 | (reference) | |

Table S6. Clinical Outcomes Comparing Switching to Apixaban (N=106) with Continuing Rivaroxaban (N=266) Among Patients Who Received Rivaroxaban Before Ischemic Stroke

Abbreviation: aHR, adjusted hazard ratio.

* Weighted incidence rates were calculated as the sum of weighted event counts divided by the sum of weighted person-months.

† Adjusted hazard ratios (aHRs) were estimated using Cox proportional hazards regression with propensity score–based overlap weighting. The overlap weights were derived from a propensity score model accounting for age, sex, alcohol drinker, congestive heart failure, hypertension, diabetes mellitus, coronary artery disease, peripheral artery disease, venous thromboembolism, chronic liver disease, anemia, hyperlipidemia, concomitant statin, NSAID, proton pump inhibitor, CYP/P-gp modulator use, and antiplatelet use before and after the index stroke.

‡ HR was not estimated because no outcome events occurred in switching to rivaroxaban group.

| Outcome | Exposure | Number of events | Person-months | Median follow-up month | Incidence rate (per 1000 person-months) | | aHR (95% CI) | P value |
|---|---|---|---|---|---|---|---|---|
| | | | | | unweighted | weighted | | |
| Recurrent ischemic stroke | Switched to apixaban | 3 | 1596.8 | 11.0 | 1.9 | 2.0 | 0.89 (0.24-3.39) | 0.87 |
| | Continued rivaroxaban | 8 | 4179.8 | 11.3 | 1.9 | 2.1 | (reference) | |
| Major bleeding | Switched to apixaban | 3 | 1638.5 | 12.1 | 1.8 | 2.0 | 0.90 (0.25-3.20) | 0.86 |
| | Continued rivaroxaban | 10 | 4165.0 | 10.8 | 2.4 | 2.2 | (reference) | |
| Composite outcome | Switched to apixaban | 6 | 1584.1 | 11.0 | 3.8 | 4.0 | 0.89 (0.36-2.24) | 0.81 |
| | Continued rivaroxaban | 18 | 4096.5 | 10.7 | 4.4 | 4.3 | (reference) | |

Figure S1. Kaplan-Meier Curves of (A) Recurrent Ischemic Stroke, (B) Major Bleeding, (C) Composite Outcome of Recurrent Ischemic Stroke and Major Bleeding Comparing Switching to a Different DOAC (DOAC-switched) with Continuing the Same DOAC (DOAC-continued) After Ischemic Stroke

Abbreviation: DOAC, direct anticoagulant.

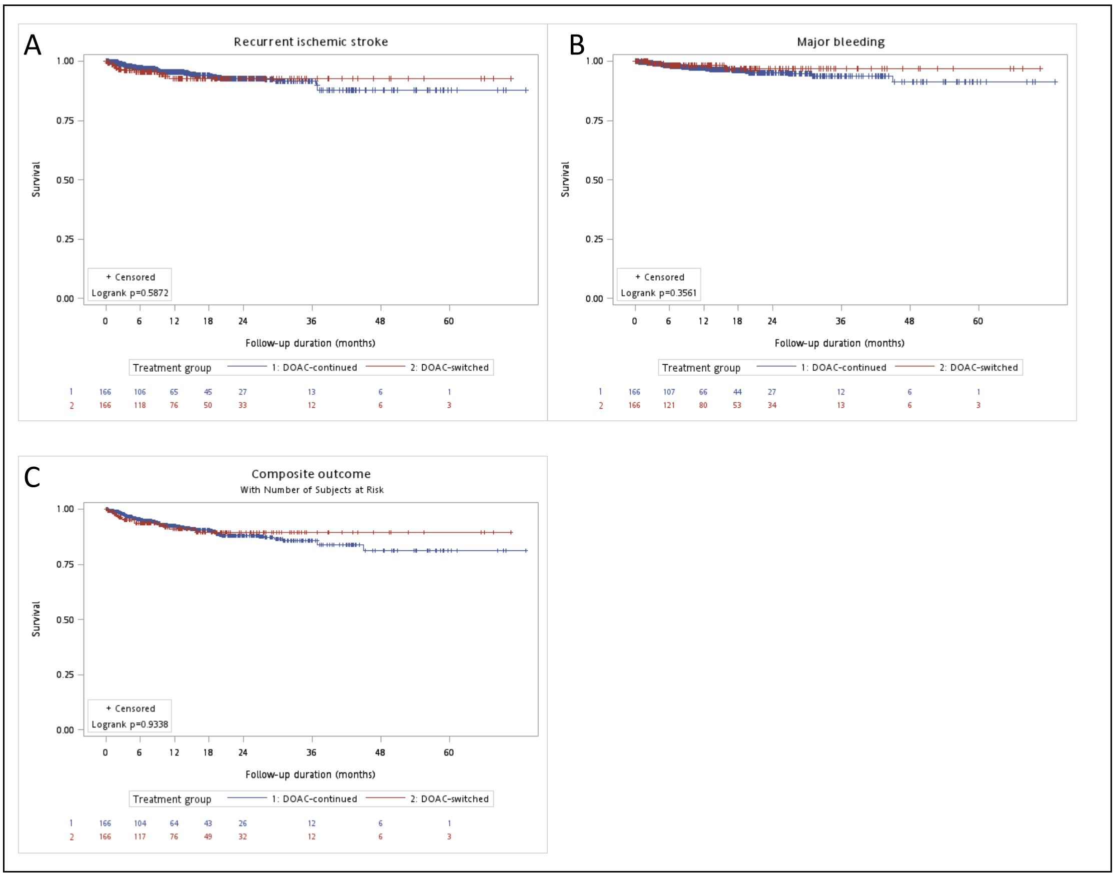

Figure S2. Kaplan–Meier Curves of (A) Recurrent Ischemic Stroke, (B) Major Bleeding, (C) Composite Outcome of Recurrent Ischemic Stroke and Major Bleeding Comparing Switching to a DOAC with a Different Mechanism of Action with Continuing a DOAC with the Same Mechanism of Action After Ischemic Stroke

Abbreviation: DOAC, direct anticoagulant.

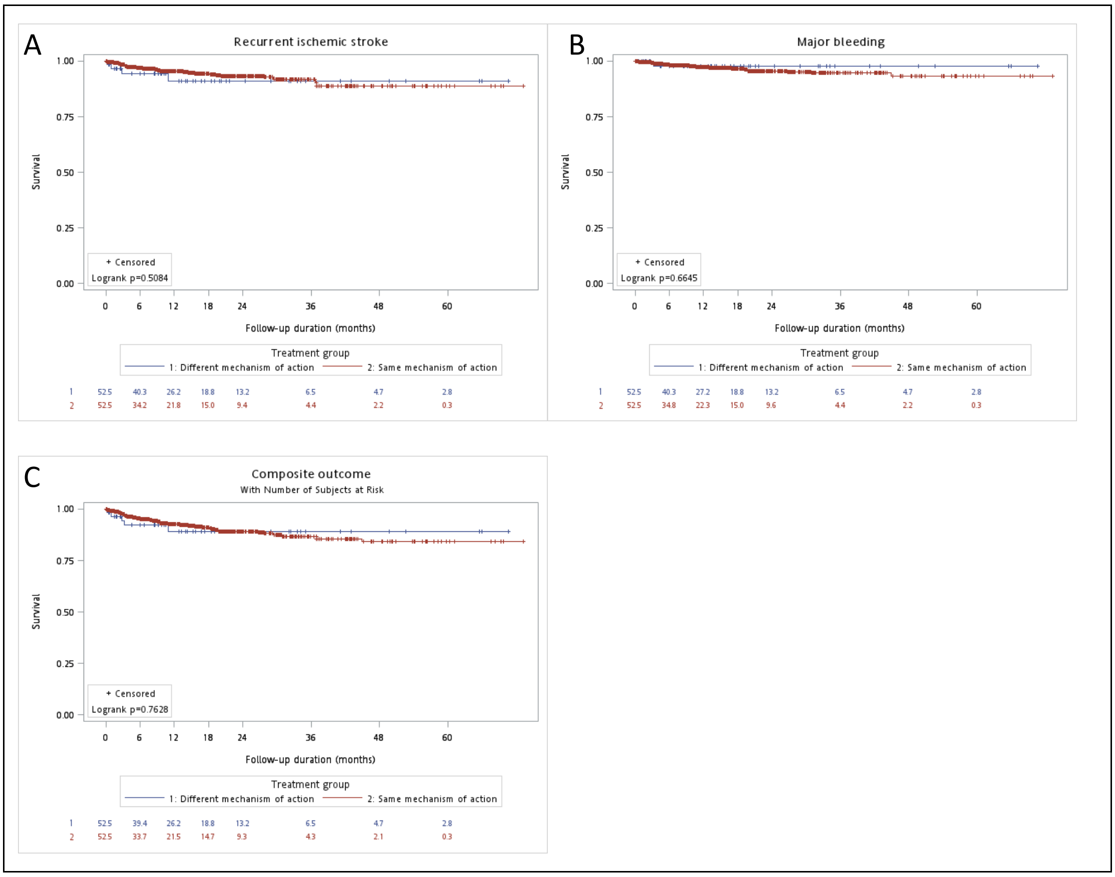

Figure S3. Kaplan–Meier Curves of (A) Recurrent Ischemic Stroke, (B) Major Bleeding, (C) Composite Outcome of Recurrent Ischemic Stroke and Major Bleeding Comparing Addition of an Antiplatelet with No Antiplatelet Addition Among Patients Who Continued the Same DOAC After Ischemic Stroke and Had No Prior Antiplatelet Therapy

Abbreviation: DOAC, direct anticoagulant.

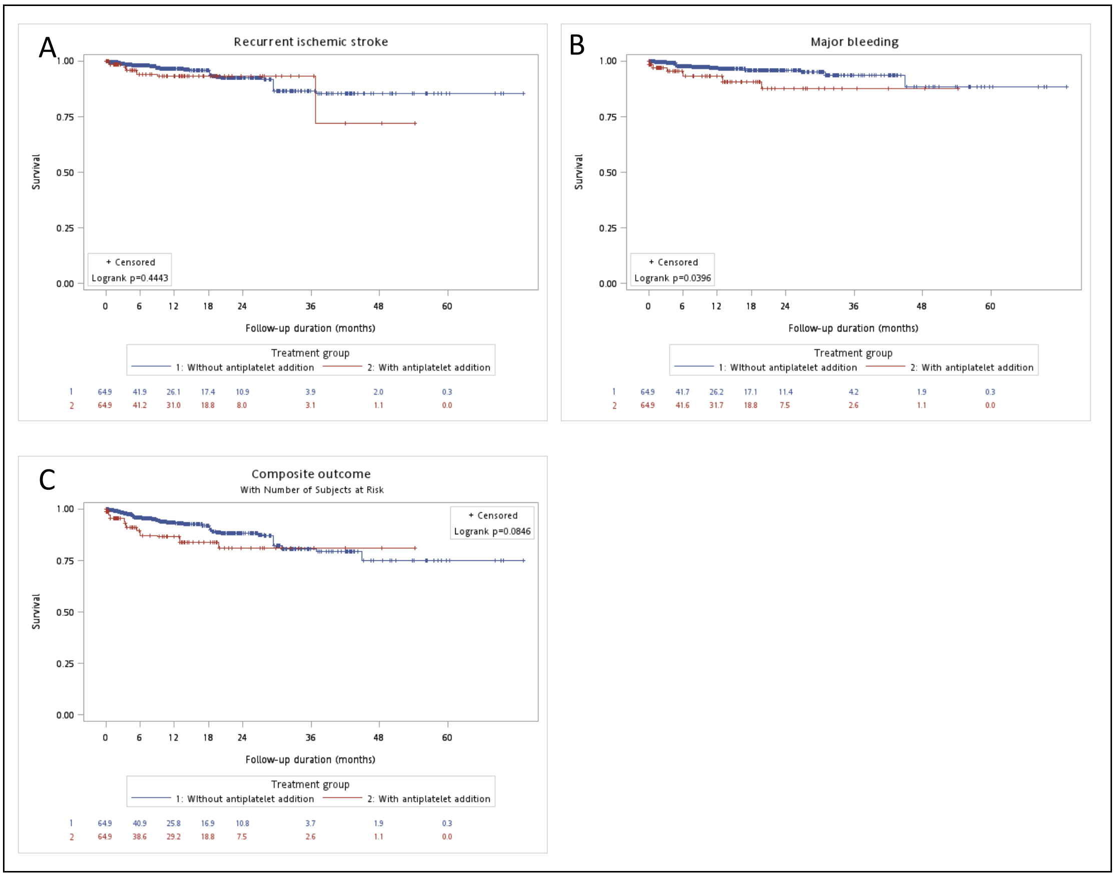

Figure S4. Kaplan–Meier Curves of (A) Recurrent Ischemic Stroke, (B) Composite Outcome of Recurrent Ischemic Stroke and Major Bleeding Comparing Switching to Rivaroxaban with Continuing Apixaban Among Patients Who Received Apixaban Before Ischemic Stroke

Abbreviation: DOAC, direct anticoagulant.

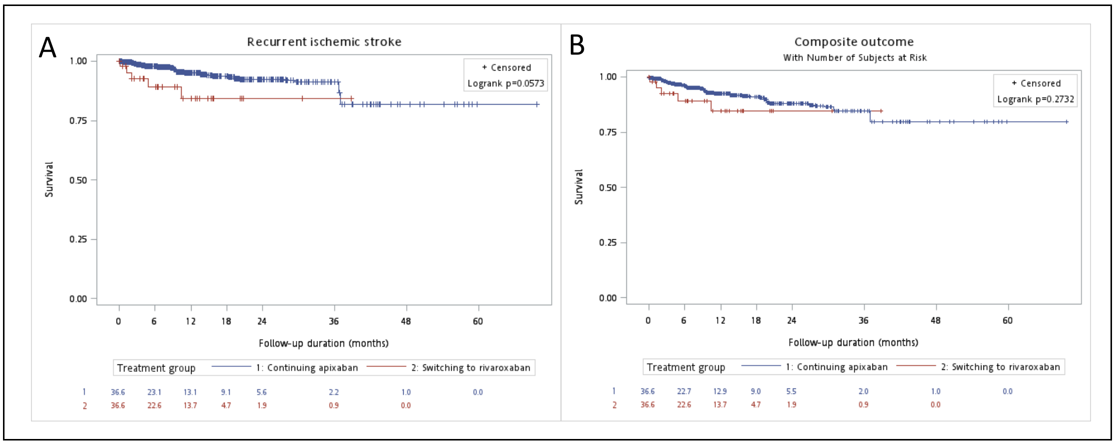

Figure S5. Kaplan–Meier Curves of (A) Recurrent Ischemic Stroke, (B) Major Bleeding, (C) Composite Outcome of Recurrent Ischemic Stroke and Major Bleeding Comparing Switching to Apixaban (N=106) with Continuing Rivaroxaban (N=266) Among Patients Who Received Rivaroxaban Before Ischemic Stroke

Abbreviation: DOAC, direct anticoagulant.

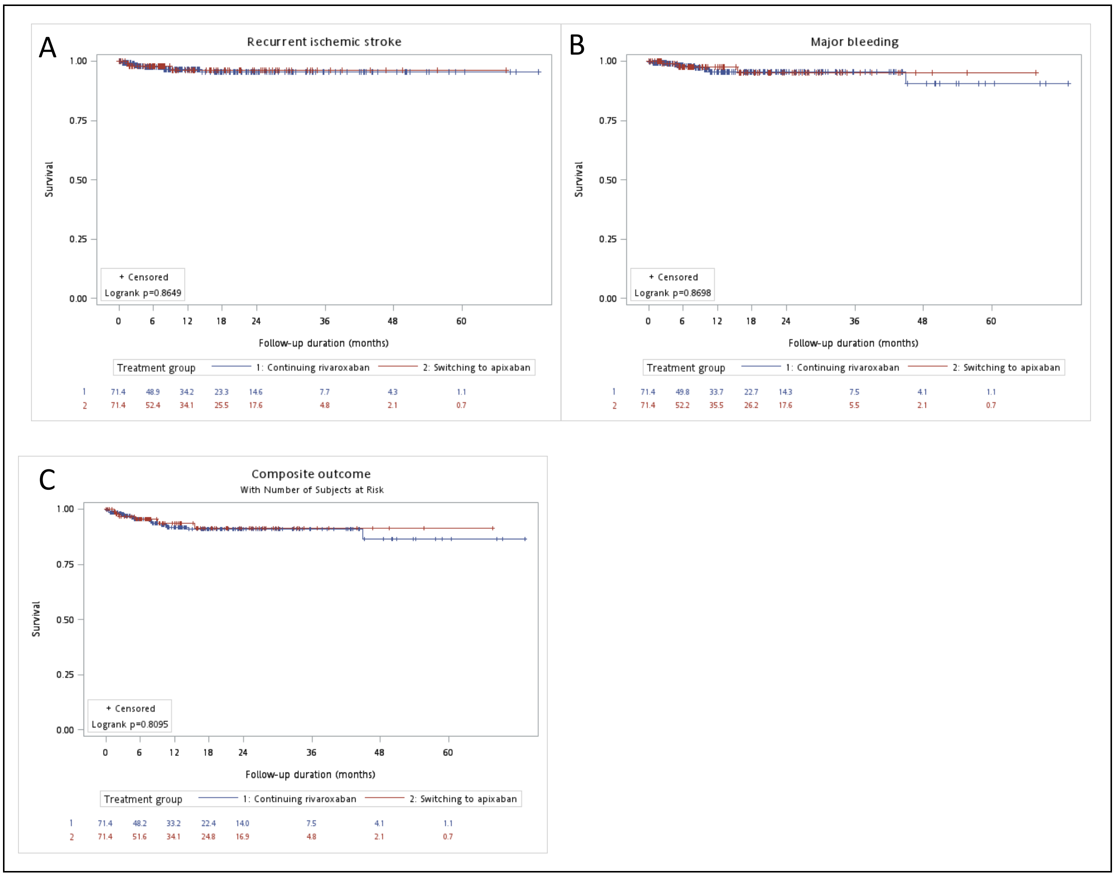
